# Postpartum Tubal Sterilization in Sickle Cell Disease in the 2012-2019 National Inpatient Sample

**DOI:** 10.1101/2024.09.30.24314263

**Authors:** Amy Luo, Alison Gemmill, Jayla L Scott, Anne E. Burke, Lydia H. Pecker

## Abstract

**Importance:** Sickle cell disease (SCD) is associated with high-risk pregnancy and low rates of hormonal contraception use. Intersectional vulnerabilities among individuals with SCD in the United States (US) raise historically and socially contingent questions about tubal sterilization in individuals with SCD. However immediate postpartum tubal sterilization (TS) rates among individuals with SCD in the US are unknown.

**Objective:** To compare rates of TS in deliveries to people with SCD and without SCD, and to determine the modifying effect of severe maternal morbidity (SMM) on the odds of TS.

**Design, Setting, and Participants:** This repeated cross-sectional study used the 2012-2019 National Inpatient Sample to estimate the rate of TS among delivery hospitalizations to people with SCD, without SCD (non-SCD), Black people with and without SCD, and people with cystic fibrosis (CF). Logistic regression models estimated the odds of TS between SCD and non-SCD deliveries, SCD and non-SCD deliveries with Black race, and SCD and CF deliveries. We examined whether SMM modified the association between TS and SCD in interaction analyses.

**Exposure:** SCD, CF

**Results:** Among 29,822,518 deliveries, 6.7% underwent TS. Among 18,860 SCD deliveries, 8.8% underwent TS. Among 2,945 CF deliveries, 6.6% underwent TS. After adjusting for patient and hospital characteristics, SCD had higher odds of TS compared to non-SCD deliveries (aOR= 1.38 [1.06,1.79]) and in a stratified analysis of deliveries coded with Black race (aOR= 1.42 [1.06,1.90]). After adjusting for patient and hospital characteristics, there was no difference in the odds of TS between SCD or CF deliveries (aOR=1.0 [0.51,2.24]). SMM more than doubled the odds of TS in SCD deliveries (interaction: aOR=2.34 [1.57,3.47]; aOR= 2.14 [1.40,3.24] in deliveries coded with Black race).

**Conclusion:** Even after accounting for patient and hospital characteristics, people with SCD have higher odds of immediate postpartum TS compared to control groups. SMM at delivery increased the odds of TS in SCD compared to all non-SCD deliveries and compared to CF deliveries. Possibly SMM severity, patient preference, or clinician recommendations inform this finding. SMM is 3-7 times more common in SCD than non-SCD pregnancies and may be a modifiable risk factor for TS in SCD deliveries.

**KEY POINTS:** 

**Question:** What are the national rates and associations of postpartum tubal sterilization (TS) among deliveries to people with sickle cell disease (SCD) compared to deliveries to people without SCD (non-SCD) and cystic fibrosis (CF)?

**Findings:** From 2012-2019, the TS rate was significantly higher in SCD deliveries (88 per 1,000 deliveries) vs non-SCD (67 per 1,000 deliveries) and CF deliveries (66 per 1,000 deliveries). SCD was positively associated with TS. SMM modified the association between TS and SCD by two-fold.

**Meaning:** Postpartum TS rate in people with SCD is high; SMM during SCD deliveries significantly increases the odds of postpartum TS.

## INTRODUCTION

Female tubal sterilization (TS) is the most common form of contraception in the United States (US) and is one of the most frequently performed surgical operations nationwide.^1,2^ TS is a nonhormonal, highly effective and irreversible form of contraception typically offered to people who have completed childbearing. In the US, women from racial minority groups, those with lower education level, lower income, disability, and those who are incarcerated have reported high rates of TS compared to the general population.^3–5^ They are also those targeted by coercive sterilization policies and practices, disproportionately receive misinformation about TS, face barriers in obtaining TS, and experience post-procedure regret.^6–11^ Individuals with chronic conditions and genetic conditions face particular historic and contemporary vulnerabilities to TS, limited studies examine TS rates in people with chronic health conditions.^12,13^

Sickle cell disease (SCD) is a genetic blood disorder that affects approximately 100,000 Americans, most of whom identify as Black. Historically, women with SCD were counseled against pregnancy.^16,17^ In a recent study, up to 44% of women with SCD were advised by their clinician they should/could not have children due to their SCD.^18,19^ Whether clinicians differentially offer or perform TS for individuals with SCD is unknown, and such recommendations might affect the timing and use of TS among individuals with SCD. Single center studies suggest that 4-30% of women with SCD use irreversible contraception.^19,20^ Despite longstanding community concerns about reproductive coercion for individuals who carry sickle cell trait and those living with SCD, little is known about TS in SCD.^14,15^

Indeed, SCD is a plausible risk factor for increased risk of undergoing TS. First, SCD is associated with three to seven times increased risk of severe maternal morbidity (SMM) and 11 to 50 times increased risk for maternal mortality.^21–23^ Experiencing or having a risk for SMM is associated with preferentially requesting TS.^24^ Second, individuals with SCD primarily identify as Black and the majority rely on public insurance – both characteristics are associated with TS use.^3,25,26^ Third, uncertainties regarding the safety of hormonal contraception in SCD may influence recommendations or preference for TS.^27^ Finally, SCD is a genetic condition associated with high risk pregnancy. This reality may affect patient and clinician perceptions of whether pregnancy, or avoiding pregnancy altogether, is advisable.^28–30^ Whether complex dynamics may lead to differential recommendation for and/or acceptance of TS among individuals with SCD is not established.

The primary objectives of this study were to: (1) estimate national rates of immediate postpartum TS in deliveries to people with and without SCD; and (2) compare the odds of TS in deliveries to people with SCD to those without SCD, to people with Black race with SCD and without, and to people with SCD to people with cystic fibrosis (CF), an autosomal recessive disease that predominately affects people who identify as White and is also associated with high risk pregnancy. Secondary objectives were to examine whether SMM at delivery modifies the odds of TS in the study groups.

## MATERIALS AND METHODS

### Data set

This retrospective study utilized data from the nationally representative 2012-2019 National Inpatient Sample (NIS). Sponsored by the Agency for Healthcare Research and Quality, the NIS is the largest available dataset comprising all-payer, inpatient discharges in the US, covering 97% of the US population.^31^

### Sample Inclusion Criteria

The analytical sample included live birth and stillbirth delivery hospitalizations between 2012 to 2019 to individuals aged 12 to 55 years at the time of delivery (**Figure S1**). Delivery hospitalizations were identified as admissions with at least one diagnosis or procedural code for vaginal or cesarean birth using *International Classification of Disease, Ninth Revision, Clinical Modification and Procedural Coding System* (ICD-9-CM/PCS) codes from January 2012 through September 2015 and ICD-10-CM/PCS codes from October 2015 through December 2019 (**Table S1**).^32,33^ Deliveries with codes for abortion, miscarriage, and inconsistent or invalid diagnostic or procedural codes were excluded.

### Delivery Analysis Groups

We defined five analytic groups: 1) SCD deliveries; 2) non-SCD deliveries; 3) SCD deliveries coded with Black race; 4) non-SCD deliveries coded with Black race; and 5) CF deliveries. SCD was identified as deliveries that contained at least one SCD diagnosis code and no sickle cell trait code. This approach has a greater than 90% positive predictive probability for identifying cases of SCD using ICD codes.^34,35^ CF was identified as deliveries with one or more CF ICD code.^36,37^ This approach is consistent with previous NIS CF studies.^36,38^ We excluded deliveries among people with both SCD and CF (N<10 unweighted deliveries).

### Outcomes

The primary outcome was immediate postpartum TS, defined as deliveries that included at least one ICD code for bilateral tubal ligation, bilateral partial salpingectomy, or bilateral total salpingectomy (**Table S1**). We excluded TS cases that contained codes for both TS and IUD initiation, TS and contraceptive implant insertion, or TS and hysterectomy (comprising <6% of cases). A sensitivity analysis including these cases found no difference in results.

The secondary outcome, intrapartum SMM, was defined as deliveries that included one or more of a set of previously validated ICD search criteria for SMM (**Table S1**).^39–41^ We used 18 of the 21 conditions defined by the Centers for Disease Control and Prevention, excluding blood transfusion, vaso-occlusive crises, and hysterectomy. These were excluded because blood transfusion has poor positive predictivity for SMM and SCD is associated with blood transfusion in pregnancy and vaso-occlusive crises.^40,42,43^ Hysterectomy was excluded because it is a procedure that also results in sterilization. A sensitivity analysis including vaso-occlusive crises in SMM identified no difference in results.

### Statistical Analysis

We compared the distribution of patient and hospital characteristics using chi-square tests, t-tests, or Wilcoxon sign rank tests where appropriate. The rate of TS was estimated as the cumulative incidence of TS per 1,000 deliveries and compared using absolute risk difference (RD).

First, we compared the characteristics and TS rates in SCD deliveries to non-SCD deliveries. Next, we performed a stratified analysis among deliveries coded with Black race and compared the characteristics and TS rates in SCD and non-SCD deliveries. This approach reduces confounding due to effects of structural and interpersonal racism on pregnancy outcomes among people with Black race.^41–43^ Last, we compared the characteristics and TS rates among SCD to CF deliveries.

We used logistic regression models to estimate odds ratios (ORs) and 95% confidence intervals (95% CIs). We estimated sequential multivariate models, incorporating covariates using forward stepwise selection and based on theoretical or subject-matter knowledge. Model fit was assessed using the Archer and Lemeshow F-adjusted mean residual test for survey sample data.^44^ To test whether SMM differentially modified the odds of TS by SCD, we assessed the interaction between SCD and SMM. The final multivariate model adjusted for maternal age, SMM, delivery mode, insurance type, median household income by zip code, hospital location and teaching status, hospital census division, year, the interaction between SCD and SMM, and the interaction between SCD and delivery mode.

Separate multivariate models were constructed to compare analytic groups: SCD vs non-SCD deliveries, SCD vs non-SCD deliveries with Black race, and SCD vs CF deliveries. To examine geographic differences in the association between TS and SCD, we repeated multivariate models among deliveries in each hospital census region and division.

We conducted all analyses using design-based analysis (Taylor linearization, and Rao and Scott’s second-order corrected Pearson statistics) to account for the NIS sampling design, following NIS-provided guidance and protocols.^45–47^ All analyses were conducted in STATA BE version 17 (StataCorp LLC, College Station, TX). Statistical significance was defined at a two-sided p-value of <0.05.

### Research Ethics and Reporting

This study received approval from the Johns Hopkins Medicine Institutional Review Board. All NIS hospital admission data are fully deidentified, so informed consent was not required. Reporting adhered to the Strengthening the Reporting of Observational Studies in Epidemiology (STROBE) guidelines.^48^ Data analysis was conducted from May to September 2024.

## RESULTS

### Patient and Hospital Characteristics of Deliveries

From 2012 to 2019, there were 29,822,518 (weighted) inpatient deliveries among people aged 12 to 55 years in the US. Among 18,860 (6.3%) SCD deliveries, 15,200 (82.5%) were deliveries with Black race. Among 2,945 CF deliveries, 2,130 (76.1%) were deliveries with White race.

The demographic and hospital characteristics of deliveries are shown (**Table 1**). SCD deliveries had higher rates of SMM and longer average length of stay compared to non-SCD deliveries.^21^ Compared to non-SCD deliveries, SCD deliveries were younger, more likely to be publicly insured, and resided in zip codes with median household incomes in lower quartiles. SCD deliveries were more likely to occur at large hospitals, urban teaching hospitals, government-owned hospitals; and most occurred in the South Atlantic, mid-Atlantic and East North Central regions. In a stratified analysis limited to deliveries coded with Black race, similar differences in were observed except that there were no between-group differences in maternal age, household income by zip code, or hospital region.

**Table 1.** Patient and hospital characteristics of delivery admissions by comparison group.

|  | **SCD**<br>N (%) | **Non-SCD**<br>N (%) | **Black, SCD**<br>N (%) | **Black, non-SCD**<br>N (%) | **CF**<br>N (%) |
| --- | --- | --- | --- | --- | --- |
| **Total admissions** |  |  |  |  |  |
| Unweighted | 3,772 | 5,960,734 | 3,040 | 831,540 | 589 |
| Weighted | 18,860 | 29,803,658 | 15,200 | 4,157,700 | 2,945 |
| **Characteristics** |  |  |  |  |  |
| Age, mean (SD), years | 27.4 (5.85) | 28.6 (5.89) | 27.1 (5.74) | 27.2 (6.05) | 28.3 (5.64) |
| **Race** |  |  |  |  |  |
| Black | 15,200 (82.5) | 4,157,870 (21.0) | 15,200 (100) | 4,157,700 (100) | 190 (76.1) |
| Hispanic | 1,285 (7.0) | 5,865,503 (29.6) | – | – | 345 (6.8) |
| White | 880 (4.8) | 14,958,803 (75.5) | – | – | 2130 (12.3) |
| Asian or Pacific Islander | 325 (1.8) | 1,680,749 (8.5) | – | – | DS |
| Native American | 45 (0.2) | 212,510 (1.1) | – | – | 0 (0) |
| Other | 680 (3.7) | 1,326,025 (6.7) | – | – | 90 (3.2) |
| **SMM** |  |  | – | – |  |
| Yes | 1,015 (5.4) | 183,925 (0.6) | 865 (5.7) | 37,295 (0.9) | 70 (2.4) |
| **Delivery Mode** |  |  |  |  |  |
| Vaginal | 10,300 (54.6) | 20,124,602 (67.5) | 8,070 (53.1) | 2,670,385 (64.2) | 1,995 (67.7) |
| Cesarean | 8,560 (45.4) | 9,679,056 (32.5) | 7,130 (46.9) | 1,487,315 (35.8) | 950 (32.3) |
| **Household income by zip code** |  |  |  |  |  |
| 1<sup>st</sup> quartile | 8,365 (45.1) | 8,296,652 (28.2) | 7,120 (46.8) | 2,028,440 (48.8) | 640 (22.1) |
| 2<sup>nd</sup> quartile | 4,495 (24.2) | 7,396,431 (25.1) | 3,690 (24.3) | 944,935 (22.7) | 785 (27.1) |
| 3<sup>rd</sup> quartile | 3,470 (18.7) | 7,283,302 (24.7) | 2,670 (17.6) | 709,005 (17.1) | 725 (25) |
| 4<sup>th</sup> quartile | 2,220 (12.0) | 6,456,283 (21.9) | 1,485 (9.8) | 423,795 (10.2) | 750 (25.9) |
| **Insurance Type** |  |  |  |  |  |
| Public<sup>a</sup> | 12,435 (66.0) | 13,001,226 (43.7) | 10,460 (68.8) | 2,702,775 (65.0) | 1215 (41.3) |
| Private | 5,530 (29.4) | 15,136,808 (50.9) | 4,030 (26.5) | 1,266,320 (30.5) | 1600 (54.4) |
| Self-pay | 390 (2.1) | 759,555 (2.6) | 300 (2.0) | 85,915 (2.1) | 30 (1.0) |
| No charge, Other<sup>b</sup> | 475 (2.5) | 864,375 (2.9) | 380 (2.5) | 96,020 (2.3) | 95 (3.2) |
| **Hospital Teaching Status** |  |  |  |  |  |
| Rural | 555 (2.9) | 2,904,294 (9.7) | 470 (3.1) | 246,370 (5.9) | 130 (4.4) |
| Urban |  |  |  |  |  |
| Teaching | 15,195 (80.6) | 7,954,727 (63.6) | 2,470 (16.3) | 845,510 (20.3) | 2230 (75.7) |
| Nonteaching | 3,110 (16.5) | 18,944,637 (26.7) | 12,260 (80.7) | 3,065,820 (73.7) | 585 (19.9) |
| **Hospital Volume** |  |  |  |  |  |
| Small | 2,380 (12.6) | 5,002,124 (16.8) | 1,920 (12.6) | 595,550 (14.3) | 340 (11.5) |
| Medium | 5,265 (27.9) | 9,017,209 (30.3) | 4,310 (28.4) | 1,323,195 (31.8) | 800 (27.2) |
| Large | 11,215 (59.5) | 15,784,325 (53.0) | 8,970 (59.0) | 2,239,125 (53.9) | 1805 (61.3) |
| **Hospital Ownership** |  |  |  |  |  |
| Government, nonfederal | 3,205 (17.0) | 3,546,635 (11.9) | 2,630 (17.3) | 578,155 (13.9) | 410 (13.9) |
| Private, no-profit | 13,480 (71.5) | 21,956,163 (73.7) | 10,805 (71.1) | 2,967,281 (71.4) | 2200 (74.7) |
| Private, invest-own | 2,175 (11.5) | 4,300,861 (14.4) | 1,765 (11.6) | 612,265 (14.7) | 335 (11.4) |
| **Hospital Region** |  |  |  |  |  |
| Northeast | 3,410 (18.1) | 4,777,009 (16.0) | 2,370 (15.6) | 628,030 (15.1) | 475 (16.1) |
| Midwest | 3,585 (19.0) | 6,306,635 (21.2) | 3,015 (19.8) | 807,450 (19.4) | 650 (22.1) |
| South | 10,260 (54.4) | 11,569,382 (38.8) | 8,720 (57.4) | 2,366,715 (56.9) | 1105 (37.5) |
| West | 1,605 (8.5) | 7,150,632 (24.0) | 1,095 (7.2) | 355,505 (8.6) | 715 (24.3) |
| **Hospital Census Division** |  |  |  |  |  |
| New England | 560 (3.0) | 1,162,955 (3.9) | 410 (2.7) | 102,555 (2.5) | 130 (4.4) |
| Mid-Atlantic | 2,850 (15.1) | 3,614,054 (12.1) | 1,960 (12.9) | 525,480 (12.6) | 345 (11.7) |
| East North Central | 2,875 (15.2) | 4,227,774 (14.2) | 2,485 (16.3) | 650,070 (15.6) | 425 (14.4) |
| West North Central | 710 (3.8) | 2,078,862 (7.0) | 530 (3.5) | 157,380 (3.8) | 225 (7.6) |
| South Atlantic | 6,720 (35.6) | 5,628,926(18.9) | 5,750 (37.8) | 1,439,455 (34.6) | 585 (19.9) |
| East South Central | 1,165 (6.2) | 1,819,945 (6.1) | 1,045 (6.9) | 348,410 (8.4) | 205 (7) |
| West South Central | 2,375 (12.6) | 4,120,511(13.8) | 1,925 (12.7) | 578,849 (13.9) | 315 (10.7) |
| Mountain | 330 (1.7) | 2,263,563 (7.6) | 255 (1.7) | 106,755 (2.6) | 170 (5.8) |
| Pacific | 1,275 (6.8) | 4,887,069 (16.4) | 840 (5.5) | 248,750 (6.0) | 545 (18.5) |
| **Year** |  |  |  |  |  |
| 2012 | 1,945 (10.3) | 3,761,961(12.6) | 1,545 (10.2) | 508,891 (12.2) | 315 (10.7) |
| 2013 | 1,910 (10.1) | 3,737,643 (12.5) | 1,515 (10.0) | 507,780 (12.2) | 300 (10.2) |
| 2014 | 2,120 (11.2) | 3,797,240 (12.7) | 1,680 (11.1) | 508,075 (12.2) | 320 (10.9) |
| 2015 | 2,175 (11.5) | 3,809,910 (12.8) | 1,780 (11.7) | 529,075 (12.7) | 390 (13.2) |
| 2016 | 2,565 (13.6) | 3,782,316 (12.7) | 2,105 (13.8) | 524,145 (12.6) | 410 (13.9) |
| 2017 | 2,560 (13.6) | 3,700,681 (12.4) | 2,035 (13.4) | 534,485 (12.9) | 460 (15.6) |
| 2018 | 2,930 (15.5) | 3,630,719 (12.2) | 2,350 (15.5) | 519,140 (12.5) | 390 (13.2) |
| 2019 | 2,655 (14.1) | 3,584,189 (12.0) | 2,190 (14.4) | 526,110 (12.7) | 360 (12.2) |
| **Length of Stay, medium (IQR), days** | 4.35 (5.02) | 2.65 (2.28) | 4.49 (5.12) | 2.93 (2.74) | 3.44 (4.1) |

N (%): Count (Percentage)
SD: Standard deviation
IQR: Interquartile range
SCD: Sickle Cell Disease
CF: Cystic Fibrosis
SMM: Severe Maternal Morbidity
DS: Data are suppressed as the number of deliveries with SCD and CF are excluded (N= unweighted cell size is 10 or fewer)
<sup>a</sup> "Public" includes Medicaid and Medicare
<sup>b</sup> "No charge, Other" includes Worker's Compensation, CHAMPUS, CHAMPVA, Title V, and other government programs

Compared to CF deliveries, SCD deliveries were younger, resided in zip codes with median household incomes in lower quartiles, more likely to be publicly insured, and more likely to occur at urban teaching hospitals and in the US South. CF deliveries were more likely than SCD deliveries to occur at rural or urban nonteaching hospitals and in the US West. SCD deliveries had a higher rate of SMM than CF deliveries. There was no difference in SCD and CF deliveries by hospital size.

### Tubal Sterilization Rates

The rate of TS in SCD deliveries was higher than non-SCD deliveries (88 per 1,000 SCD deliveries vs 67 per 1,000 non-SCD deliveries, RD 22 per 1,000 deliveries; p<0.001), and higher than CF deliveries (66 per 1,000 CF deliveries, RD 22 per 1,000 deliveries p<0.05) (**Table S3**). Among deliveries with Black race, the TS rate was also higher among SCD than non-SCD deliveries (92 per 1,000 SCD deliveries vs 69 per 1,000 non-SCD deliveries, RD 23 per 1,000 deliveries, p<0.001).

### Characteristics Associated with Tubal Sterilization

In all SCD deliveries and SCD deliveries with Black race, TS was associated with older age at delivery, presence of SMM, cesarean delivery, lower median household income by zip code, government hospital ownership, and delivery in the US South region (**Table S4**). Among all non-SCD deliveries and non-SCD deliveries with Black race, TS was also associated with public insurance, rural hospital location, large hospital volume, and calendar year. Among CF deliveries, TS was only associated with older age at delivery.

Multivariate models comparing SCD to non-SCD deliveries identified higher odds of TS in SCD deliveries (adjusted OR (aOR): 1.38 [1.06,1.79]; **Table 2**). Among deliveries with Black race, SCD was also associated with higher odds of TS (aOR: 1.42 [1.06,1.90]). There was no difference in the odds of TS among SCD deliveries compared to CF deliveries (OR: 1.37 [0.97,1.93]; aOR: 0.82 [0.52,2.25]).

**Table 2.** Adjusted odds of postpartum tubal sterilization.

|  | All deliveries |  | Deliveries coded with Black race |  | SCD or CF deliveries |  |
| --- | --- | --- | --- | --- | --- | --- |
|  | aOR | 95% CI | aOR | 95% CI | aOR | 95% CI |
| **Main Effects** |  |  |  |  |  |  |
| Comparison Group |  |  |  |  |  |  |
| Comparison [Reference Group] | 1.00 [non-SCD] | – | 1.00 [Black, non-SCD] | – | 1.00 [CF] | – |
| SCD | 1.38** | 1.06 - 1.79 | 1.42** | 1.06 - 1.90 | 0.82 | 0.52 - 2.25 |
| SMM |  |  |  |  |  |  |
| No | Ref | – | Ref | – | Ref | – |
| Yes | 0.75** | 0.72 - 0.78 | 0.91** | 0.84 - 0.99 | 2.25 | 0.60 - 8.46 |
| Delivery Mode |  |  |  |  |  |  |
| Vaginal | Ref | – | Ref | – | Ref | – |
| Cesarean | 8.31*** | 8.13 - 8.49 | 6.82*** | 6.60 - 7.06 | 6.93*** | 3.13 - 15.33 |
| Characteristics |  |  |  |  |  |  |
| Age, years | 1.12*** | 1.12 - 1.13 | 1.13*** | 1.12 - 1.13 | 1.12*** | 1.10 - 1.15 |
| Insurance Type |  |  |  |  |  |  |
| Public | Ref | – | Ref | – | Ref | – |
| Private | 0.50*** | 0.49 - 0.50 | 0.67*** | 0.66 - 0.69 | 0.67*** | 0.51 - 0.88 |
| Self-pay | 0.44*** | 0.42 - 0.46 | 0.33*** | 0.30 - 0.36 | 0.33** | 0.12 - 1.04 |
| No charge, Other | 0.61*** | 0.59 - 0.63 | 0.78*** | 0.73 - 0.84 | 0.89 | 0.43 - 1.62 |
| Median Household Income by Zip Code |  |  |  |  |  |  |
| 0-25<sup>th</sup> percentile | Ref | – | Ref | – | Ref | – |
| 26th-50<sup>th</sup> percentile | 0.93*** | 0.92 - 0.95 | 0.89*** | 0.85 - 0.90 | 0.76* | 0.56 - 1.03 |
| 51st-75<sup>th</sup> percentile | 0.78*** | 0.77 - 0.79 | 0.77*** | 0.72 - 0.77 | 0.75* | 0.55 - 1.04 |
| 76<sup>th</sup>-100<sup>th</sup> percentile | 0.57*** | 0.55 - 0.58 | 0.58*** | 0.53 - 0.60 | 0.40*** | 0.25 - 0.62 |
| Hospital Location and Teaching Status |  |  |  |  |  |  |
| Rural | Ref | – | Ref | – | Ref | – |
| Urban, nonteaching | 0.68*** | 0.66 - 0.70 | 0.62*** | 0.57 - 0.64 | 0.83 | 0.43 - 1.60 |
| Urban, teaching | 0.59*** | 0.58 - 0.61 | 0.58*** | 0.54 - 0.60 | 0.81 | 0.44 - 1.49 |
| Hospital Census Division |  |  |  |  |  |  |
| New England | Ref | – | Ref | – | Ref | – |
| Middle Atlantic | 0.96 | 0.90 - 1.03 | 1.22*** | 1.06 - 1.34 | 0.93 | 0.44 - 1.94 |
| East North Central | 1.21*** | 1.14 - 1.29 | 1.68*** | 1.58 - 1.98 | 1.21 | 0.57 - 2.55 |
| West North Central | 1.30*** | 1.21 - 1.39 | 1.46*** | 1.32 - 1.84 | 0.74 | 0.28 - 2.99 |
| South Atlantic | 1.41*** | 1.34 - 1.49 | 2.02*** | 1.77 - 2.21 | 1.50 | 0.76 - 2.99 |
| East South Central | 1.52*** | 1.41 - 1.62 | 2.36*** | 2.00 - 2.53 | 2.04 | 0.94 - 4.44 |
| West South Central | 1.70*** | 1.60 - 1.80 | 2.52*** | 2.12 - 2.64 | 1.94 | 0.94 - 4.03 |
| Mountain | 1.24*** | 1.17 - 1.32 | 1.49*** | 1.33 - 1.72 | 0.72 | 0.21 - 2.41 |
| Pacific | 1.08*** | 1.02 - 1.15 | 1.24*** | 1.07 - 1.36 | 1.22 | 0.56 - 2.65 |
| **Year** |  |  |  |  |  |  |
| 2012 | Ref | – | Ref | – | Ref | – |
| 2013 | 1.00 | 0.95 - 1.05 | 0.97 | 0.90 - 1.05 | 1.03 | 0.65 - 1.65 |
| 2014 | 0.98 | 0.94 - 1.03 | 0.94 | 0.87 - 1.02 | 0.77 | 0.47 - 1.26 |
| 2015 | 0.94** | 0.90 - 0.99 | 0.87*** | 0.82 - 0.95 | 0.59** | 0.35 - 0.99 |
| 2016 | 0.91*** | 0.87 - 0.96 | 0.82*** | 0.76 - 0.89 | 0.77 | 0.48 - 1.21 |
| 2017 | 0.89*** | 0.85 - 0.93 | 0.75*** | 0.71 - 0.82 | 0.72 | 0.46 - 1.15 |
| 2018 | 0.87*** | 0.83 - 0.91 | 0.74*** | 0.69 - 0.80 | 0.51*** | 0.32 - 0.81 |
| 2019 | 0.85*** | 0.81 - 0.89 | 0.71*** | 0.67 - 0.77 | 0.60** | 0.37 - 0.96 |
| **Interaction** |  |  |  |  |  |  |
| SMM by SCD | 2.34*** | 1.57 - 3.47 | 2.14*** | 1.40 - 3.25 | 0.82 | 0.21 - 3.27 |
| Cesarean delivery by SCD | 0.66** | 0.49 - 0.89 | 0.75* | 0.54 - 1.05 | 0.79 | 0.34 - 1.85 |

\*\*\* p<0.01, \*\* p<0.05, \* p<0.1
aOR: Adjusted Odds Ratio
Ref: Reference group (aOR=1.0)
95% CI: 95% Confidence Interval
SCD: Sickle Cell Disease
CF: Cystic Fibrosis
SMM: Severe Maternal Morbidity

The association between TS and SCD varied by hospital census region (**Figure 1**) and division (**Figure S1**). Increased odds of TS among SCD deliveries were observed in the US South region (aOR= 1.44 [1.04-1.99]), largely driven by the East South Central division (aOR= 3.60 [1.75,7.40]). There were no significant differences in the odds of TS among SCD deliveries in other regions and divisions.

**Figure 1.**
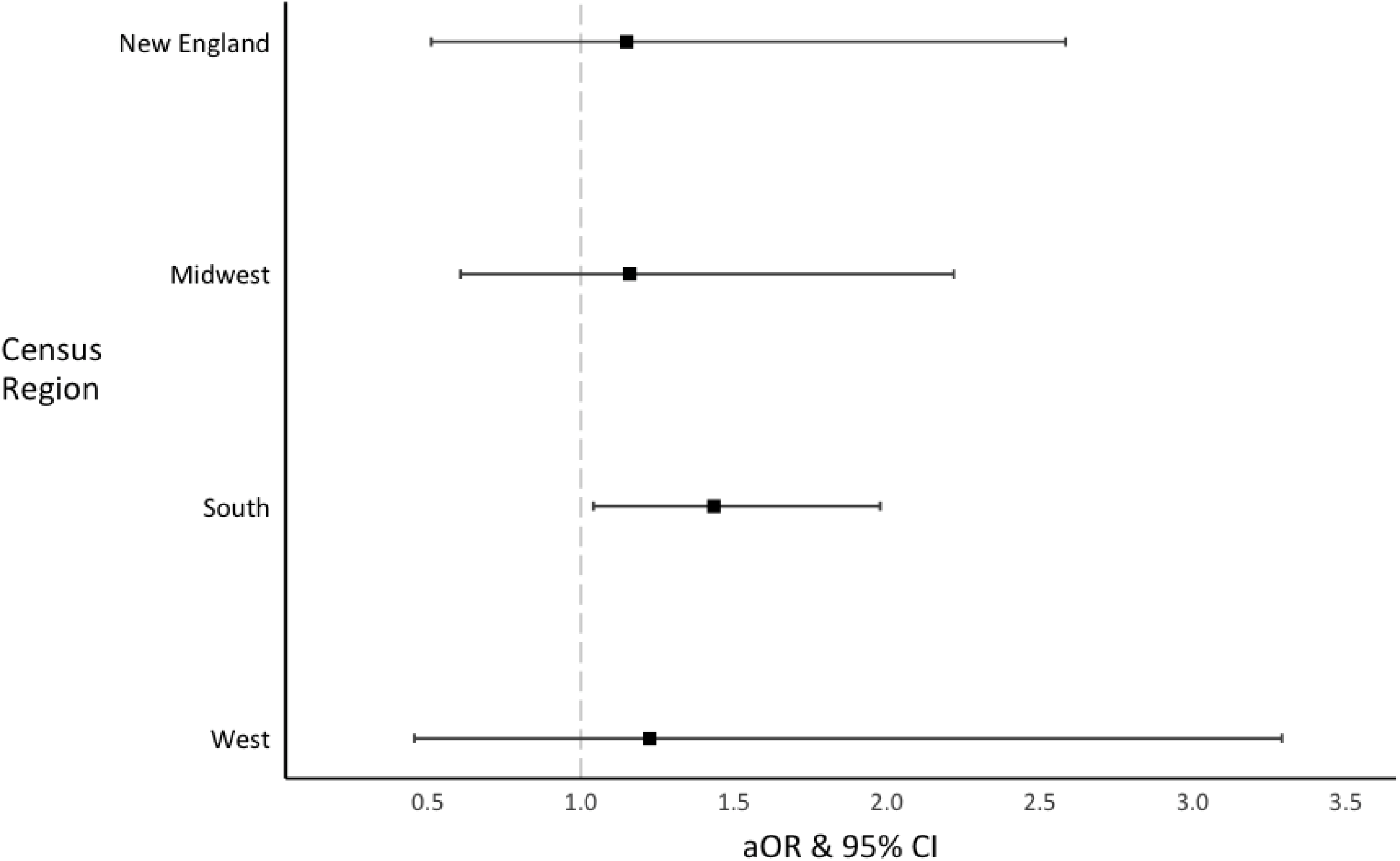
Adjusted odds of postpartum tubal sterilization among sickle cell disease deliveries compared with non-sickle cell disease deliveries, stratified by hospital census region Multivariate models adjusted for SCD, age, SMM, delivery mode, insurance type median household income by zip code, hospital location and teaching status, hospital census division, year, and the interaction of SCD and SMM, and SCD and delivery mode. aOR: Adjusted Odds Ratio 95% CI: 95 %Confidence Interval

### Effect Measure Modification by SMM

SMM modified the association between TS and SCD (**Table 2**). After adjusting for patient and hospital characteristics, SMM in SCD deliveries doubled the odds of TS compared to non-SCD deliveries (aOR: 2.34 [1.57,3.47]) and in deliveries with Black race (aOR: 2.14 [1.40,3.25]). In contrast, a sensitivity analysis of the modification of the association between TS and CF compared to deliveries without CF found no modification (aOR= 2.79 [0.64,12.09]) (**Table S5**).

## DISCUSSION

This study uses a nationally representative sample of hospital discharge data to report novel population-based estimates of rates of postpartum TS among people with SCD and CF. Between 2012-2019, the TS rate per 1,000 deliveries was 88 in people with SCD, 67 in people without SCD, and 66 in people with CF. As in the general population, delivering in the US South increased the odds of TS for people with SCD. The odds of TS among SCD deliveries more than doubled among deliveries complicated by SMM, but SMM had no effect on TS for CF deliveries. SMM risk is modifiable in SCD.^49^ The urgent need to reduce SMM in people with SCD can be coupled to future studies examining whether reducing SMM also changes postpartum TS rates for people with SCD.^50^

We hypothesized that patient or hospital characteristics contribute to the postpartum TS rates in SCD. However, even after adjusting for age, SMM, delivery mode, insurance type, household income, hospital location and teaching status, hospital census division, and year, the odds of TS remained 1.38 times higher in individuals with SCD. In the race-stratified cohort, the odds of postpartum TS were also higher among those with SCD, suggesting SCD-related factors, not solely race, drive these findings. We then hypothesized that SCD, as a genetic condition, might contribute to the increased odds of postpartum TS. To test this hypothesis, we examined postpartum TS in SCD and CF and found that, although the rate of postpartum TS is higher in SCD, the adjusted odds of postpartum TS were not different between individuals with SCD and CF. The lack of differences in adjusted odds of postpartum TS between individuals with SCD and CF suggests that the higher postpartum TS rates in individuals with SCD compared to those with CF are primarily driven by differences in patient and hospital characteristics between the groups.

Here, delivery geography influenced the odds of postpartum TS for individuals with SCD. Compared to individuals without SCD, those with SCD had higher odds of undergoing TS in the South, particularly in the East South Central division – a pattern not observed elsewhere. The reasons for geographic variation in postpartum TS are not fully elucidated. Possibly, SCD contributes to described differences in postpartum TS rates in the US.^51,52^ This is important since the US South has the highest concentrations of individuals living with SCD and inadequate numbers of SCD experts to provide care.^53^ Further, individuals with SCD who are at risk for miscarriage and receive fragmented reproductive healthcare may be disproportionately affected by the growing restrictions in access to abortion and limited access to long-acting reversible contraceptive methods in the US South.^19,51,54,55^ Overall, the lack of coordinated SCD care and presence of maternity care deserts in regions with higher concentrations of people with SCD may further contribute to higher use of TS.^19,20,53,56^

This is the first study to demonstrate a modifying effect of SMM on postpartum TS for individuals with SCD. In general, deliveries complicated by SMM have higher rates of postpartum TS and pregnant people with SCD are 5-7 times more likely to experience SMM.^21,57,58^In this study, SMM more than doubled the odds of postpartum TS among people with SCD. In contrast, SMM did not modify the odds of postpartum TS among people with CF. The effect modification of SMM on TS for SCD compared to CF requires a qualitative understanding of SMM which is inadequately appraised through the diagnosis codes used in this study. The extent to which SMM modifies TS in SCD may be a consequence of the nature of the SMM and this affects patient preferences and clinical practice.^59^ Given the high rates of SMM in this population, limits to existing contraceptive counseling and hormonal contraceptive safety concerns may influence the uptake of or recommendation for postpartum TS.^18,19,60,61^ Finally, SMM at delivery may also be a marker for pregnancy complications before delivery in individuals with SCD; a very difficult pregnancy course punctuated by SMM at delivery may shape recommendations for no future pregnancies and increase use of TS at delivery in SCD.^18,19^ Future research can help establish the extent to which SMM at delivery constitutes a modifiable risk factor for postpartum TS in SCD.^21,62^

Fifty years ago, Drs. James Bowman and Charles Whitten voiced community concerns that testing for SCD and trait might lead to coercive reproductive care for the SCD community.^14,63^ This study cannot determine whether TS rates among individuals with SCD are a manifestation of reproductive coercion, a risk that may increase as growing restrictions to lifesaving reproductive healthcare are implemented, especially in the US South where most individuals with SCD live.^55,64,65^ This is the first study to empirically appraise a national dataset on postpartum TS for individuals with SCD or CF; follow-up studies in subsequent NIS will be possible to assess trends. However, this study is not without limitations. First, estimates are limited to postpartum TS and do not capture TS procedures in outpatient settings, outside of the postpartum period, or male partner sterilization. Second, as with administrative data, findings are limited to what is reported for billing purposes during delivery admission. As such, SMM at delivery cannot capture clinical severity across pregnancy. Third, this secondary analysis cannot identify all factors that shape individual preference or clinician recommendations for postpartum TS. For instance, how sterilization decisions inform cesarean delivery mode or vice versa in people with SCD compared to their counterparts requires further study.

In conclusion, people with SCD undergo postpartum TS at higher rates than the general population, a difference unexplained by differences in patient or hospital characteristics. Experiencing SMM during delivery modified the association between TS and SCD by two-fold, suggesting that the urgently needed interventions to reduce SMM in SCD pregnancy might modify postpartum TS rates. Individuals with SCD and their clinicians balance the risks associated with pregnancy and contraception in the context of a life-limiting illness associated with disturbing and life-altering SMM rates.^19,28,59^ Research to understand these findings – including the extent to which pragmatic realities, patient preference, or structural or interpersonal bias inform TS rates – is needed. This is especially critical as states across the country eliminate or severely restrict access to abortion, opening the door to reproductive coercion for an underrepresented patient population with multiple marginalized identities.

## Data Availability

All data produced in the present study are managed by the Agency for Healthcare Research and Quality and is only available through registration with this entity and completion of their required training module. For more information on accessing HCUP data visit www.hcup-us.ahrq.gov.

https://hcup-us.ahrq.gov/nisoverview.jsp

## Additional Contributions

The authors gratefully acknowledge the patients, and the Agency for Healthcare Research and Quality and HCUP data organization partners that participated in the 2012-2019 National Inpatient Sample (https://hcup-us.ahrq.gov/partners.jsp).

## Author Contributions

AL: conception and design; analysis; interpretation; drafting and editing. AG: conception and design; interpretation; drafting and editing. JLS: editing. AEB: conception and design; drafting and editing. LHP: conception and design; interpretation; drafting and editing. All authors approved of the final version and have confirmed compliance with the journal’s requirements for authorship.

## Data Sharing Statement

Data will not be shared. Data are made available by the Agency for Healthcare Research and Quality’s Healthcare Cost and Utilization Project upon registration and completion of their required training module (www.hcup-us.ahrq.gov).

## Abbreviations List

95% CI: 95% Confidence Interval
aOR: Adjusted Odds Ratio
CF: Cystic Fibrosis
ICD: *International Classification of Disease*
ICD-10-CM/PCS: *International Classification of Disease, Tenth Revision, Clinical Modification and International Classification of Disease, Tenth Revision, Procedural Coding System*
ICD-9-CM/PCS: *International Classification of Disease, Ninth Revision, Clinical Modification and International Classification of Disease, Ninth Revision, Procedural Coding System*
NIS: National Inpatient Sample
OR: Odds Ratio
RD: Rate Difference
SCD: Sickle Cell Disease
SMM: Severe Maternal Morbidity
TS: Tubal Sterilization during the Immediate Postpartum Period
US: United States

