## Supplemental Materials for "Postpartum Tubal Sterilization in Sickle Cell Disease in the 2012-2019 National Inpatient Sample"

**APPENDIX**

**Table S1**. ICD-9-CM/PCS and ICD-10-CM/PCS codes used identify independent and dependent variables

**Table S2.** Proportion of missing data by comparison group

**Table S3.** Survey-weighted rates of postpartum tubal sterilization

**Table S4.** Association of patient and hospital characteristics with postpartum tubal sterilization by comparison group

**Table S5.** Sensitivity Analysis: Effect measure modification of the association of postpartum tubal sterilization and cystic fibrosis and by severe maternal morbidity

**Figure S1.** Flow diagram of study analytical sample inclusion

**Figure S2.** Adjusted odds of postpartum tubal sterilization among sickle cell disease deliveries compared with non-sickle cell disease deliveries, stratified by hospital census division

**Table S1.** *ICD-9-CM/PCS* and *ICD-10-CM/PCS* codes used identify independent and dependent variables

|  | **ICD-9** | | **ICD-10** | |
| --- | --- | --- | --- | --- |
| **Inclusion Criteria** | **DX** | **PR** | **DX** | **PR** |
| Outcome of delivery | V27*  650  651.01  651.11  651.21 |  | Z37* |  |
| Normal, uncomplicated delivery |  |  | O80* |  |
| Complicated delivery |  |  | O60.1*  O60.2*  O61*-O77* |  |
| Cesarean birth | 669.7* | 74.0  74.1  74.2  74.4  74.99 | O82* | 10D00Z0  10D00Z1  10D00Z2 |
| Forceps, vacuum, and breech extraction, Assisted delivery, Episiotomy |  | 72*  73.22  73.59  73.6 |  | 10D07Z3  10D07Z4  10D07Z5  10D07Z6  10D07Z7  10D07Z8  10E0XZZ  10S07ZZ  0W8NXZZ |
| **Exclusion Criteria** | **DX** | **PR** | **DX** | **PR** |
| Ectopic or molar pregnancy | 630*  631*  632*  633* | 74.3  66.62 | O00*  O01* | 10T20ZZ  10T23ZZ  10T24ZZ  10T27ZZ  10T28ZZ |
| Pregnancy with abortive outcome or abortion | 634*  635*  636*  637*  638*  639* | 69.01  69.51  74.91  75.0 | O02*  O03*  O04*  O05*  O06*  O07*  O08*  O36.4*  O33.2* | 10A0* |
| **Exposures** | **DX** | **PR** | **DX** | **PR** |
| Sickle Cell Disease | 282.6*  282.41  282.42 |  | D57.0*  D57.1*  D57.2*  D57.4* |  |
| Sickle Cell Trait | 282.5 |  | D57.3 |  |
| Cystic Fibrosis | 277.0* |  | E84.* |  |
| **Pregnancy Outcomes** | **DX** | **PR** | **DX** | **PR** |
| Bilateral Tubal Ligation, Bilateral Total or Partial Salpingectomy | V25.2 | 66.2  66.3*  66.5*  66.63 | Z30.2 | 0U57*  0UL7*  0UF7*  0UB7*  0UT7* |
| Intrauterine Device | V25.11 |  | Z30.014  Z30.430 | 0UH90HZ 0UH97HZ 0UH98HZ 0UHC7HZ 0UHC8HZ |
| Subdermal Implant Contraceptive | V25.5 |  | Z30.017 | 0JHF3HZ 0JHH3HZ 0JHG3HZ |
| Hysterectomy |  | 68.3*  68.4*  68.5*  68.6*  68.7*  68.8*  68.9* |  | 0UB90ZZ  0UB93ZZ  0UB94ZZ  0UB97ZZ  0UB98ZZ  0UT90Z* 0UT94Z* 0UT97Z* 0UT98Z* 0UT9FZ* |
| **Severe Maternal Mortality** | **DX** | **PR** | **DX** | **PR** |
| Acute Myocardial Infarction | 410.* |  | I21.**  I22.* |  |
| Aneurysm | 441.* |  | I71.**  I79.0 |  |
| Acute Renal Failure | 584.*  669.3* |  | N17.*  O90.4 |  |
| Adult Respiratory Distress Syndrome | 518.5* 518.81 518.82 518.84 799.1 |  | J80  J95.1  J95.2  J95.3  J95.82*  J96.0*  J96.2*  J96.9*  R06.03  R09.2 |  |
| Amniotic Fluid Embolism | 673.1* |  | O88.112  O88.113  O88.119  O88.12  O88.13 |  |
| Cardiac Arrest / Ventricular Fibrillation | 427.41 417.42 427.5 |  | I46.*  I49.0* |  |
| Conversion of Cardiac Rhythm |  | 99.6* |  | 5A2204Z  5A12012 |
| Disseminated Intravascular Coagulation | 286.6  286.9 641.3*  666.3* |  | D65  D68.8  D68.9  O45.002  O45.003 O45.009  O45.012 O45.013 O45.019  O45.022 O45.023 O45.029  O45.092 O45.093 O45.099  O46.002 O46.003 O46.009  O46.012 O46.013 O46.019  O46.022 O46.023 O46.029  O46.092 O46.093 O46.099  O67.0  O72.3 |  |
| Eclampsia | 642.6* |  | O15.* |  |
| (Acute) Heart Failure / Arrest during Surgery or Procedure | 997.1 |  | I97.12*  I97.13*  I97.71* |  |
| Puerperal Cerebrovascular Disorders | 362.34  430  431  432  433  434  435  436  437*  671.5* 674.0* 997.02 |  | G45.*  G46.*  H34.0*  I60  I61  I62  I63  I64  I65  I66  I67  I68.*  O22.50  O22.52  O22.53  I97.81*  I97.82*  O87.3 |  |
| Pulmonary Edema / Acute Heart Failure | 518.4 428.0 428.1  428.20  428.21  428.23  428.30  428.31  428.33  428.40  428.41  428.43  428.9 |  | J81.0  I50.1  I50.20  I50.21  I50.23  I50.30  I50.31  I50.33  I50.40  I50.41  I50.43  I50.810  I50.811  I50.813  I50.814  I50.82  I50.83  I50.84  I50.89  I50.9 |  |
| Severe Anesthesia Complications | 668.0*  668.1*  668.2*  995.4  995.86 |  | O29.112  O29.113 O29.119  O29.122  O29.123 O29.129  O29.192 O29.193 O29.199  O29.212 O29.213 O29.219  O29.292 O29.293 O29.299  O74.0  O74.1  O74.2  O74.3  O89.0*  O89.1  O89.2  T88.2**A  T88.3**A |  |
| Sepsis | 038.*  670.2*  998.02  995.91  995.92  785.52  449 |  | O85  R65.21  R65.20  T81.44*A  T81.12*A  I76  O86.04  A40.*  A41.*  A32.7 |  |
| Shock | 669.1* 785.50  785.51  785.59  995.0  998.0*  995.4 |  | O75.1  R57.*  T78.2**A  T88.2**A  T88.6**A  T81.10*A  T81.11*A  T81.19*A |  |
| Air and Thrombotic Embolism | 415.1*  673.0*  673.2*  673.3*  673.8* |  | I26.*  O88.012  O88.013  O88.019  O88.02  O88.03  O88.112  O88.113  O88.119  O88.12  O88.13  O88.212  O88.213  O88.219  O88.22  O88.23  O88.312  O88.313  O88.319  O88.32  O88.33  O88.812  O88.813  O88.819  O88.82  O88.83  T80.0**A |  |
| Temporary Tracheostomy |  | 31.1 |  | 0B110F4  0B113F4  0B114F4  0B110Z4  0B113Z4  0B114Z4 |
| Ventilation |  | 96.70  96.71  96.72  93.90  96.01  96.02  96.03  96.05 |  | 5A1935Z  5A1945Z  5A1955Z |

Severe Maternal Morbidity, excluding Sickle Cell Disease with crisis, Hysterectomy, and Blood Transfusion

* Includes all codes that have additional digits, as long as digits preceding the asterisk match

ICD-9-CM/PCS: *International Classification of Diseases, Ninth Revisions, Clinical Modification and Procedural Coding System*

ICD-10-CM/PCS: *International Classification of Diseases, Tenth Revisions, Clinical Modification and Procedural Coding System*

**Table S2.** Proportion of missing data by comparison group

| **Variable** | **SCD** | **Non-SCD** | **Black, SCD** | **Black, Non-SCD** | **CF** |
| --- | --- | --- | --- | --- | --- |
| Race | 2.4% | 5.4% | – | – | 4.9% |
| Income quartile by zip code | 1.6% | 1.2% | 1.5% | 1.2% | 1.5% |
| Insurance type | 0.2% | 0.1% | 0.2% | 0.2% | 0.2% |

SCD: Sickle Cell Disease

CF: Cystic Fibrosis

**Table S3.** Survey-weighted rates of postpartum tubal sterilization

|  | **All deliveries** | **SCD** | **Non-SCD** | **Black, SCD** | **Black, non-SCD** | **CF** |
| --- | --- | --- | --- | --- | --- | --- |
| **Delivery Admissions (weighted)** | 29,822,518 | 18,860 | 29,803,658 | 15,200 | 4,157,700 | 2,945 |
| **Overall tubal sterilization rate** | 6.7% | 8.8% | 6.7% | 9.2% | 6.5% | 6.6% |
| **SCD Status** | | | | | | |
| SCD | 8.8% | – | – | – | – | – |
| Non-SCD | 6.7% | – | – | – | – | – |
| *Difference, SCD - non-SCD* | 2.2%*** |  |  | – | – | – |
| **Race** | | | | | | |
| White | 6.3% | 5.7% | 6.3% | – | – | 6.8% |
| Black | 6.9% | 9.2% | 6.9% | – | – | 7.9% |
| Hispanic | 8.5% | 11.7% | 8.5% | – | – | 7.2% |
| Asian or Pacific Islander | 4.2% | DS | 4.2% | – | – | 0.0% |
| Native American | 7.0% | 0.0% | 7.0% | – | – | – |
| Other | 5.7% | 5.9% | 5.7% | – | – | 5.6% |
| *Difference, Black - Hispanic* | -1.6%*** | -2.0% | -1.6% | – | – | 0.6% |
| *Difference, Black - White* | 0.6%*** | 3.2%* | 0.6%*** | – | – | 1.1% |
| **SMM** | | | | | | |
| Yes | 8.7% | 16.7% | 8.6% | 17.9% | 10.5% | DS |
| No | 6.6% | 8.4% | 6.6% | 8.7% | 6.9% | 6.4% |
| *Difference, Yes - No* | 2.0%*** | 8.4%*** | 2.0%*** | 9.3%*** | 3.6%*** | 7.9%*** |
| **Delivery Mode** | | | | | | |
| Vaginal | 2.1% | 3.0% | 2.1% | 3.1% | 3.0% | 2.3% |
| Cesarean | 16.2% | 15.9% | 16.2% | 16.1% | 15.2% | 15.8% |
| *Difference, Cesarean - Vaginal* | 14.1%*** | 12.9%*** | 14.1%*** | 13.0%*** | 12.9%*** | 13.5% |
| **Insurance Type** | | | | | | |
| Public | 8.1% | 9.3% | 8.1% | 9.5% | 6.6% | 8.2% |
| Private | 5.6% | 8.0% | 5.6% | 8.6% | 6.6% | 5.3% |
| Self-pay | 4.9% | 5.1% | 4.9% | DS | 3.3% | 0.0% |
| No charge, Other | 5.7% | 8.4% | 5.7% | DS | 6.4% | DS |
| **Median Household Income by Zip Code** | | | | | | |
| 0-25th percentile | 8.0% | 9.4% | 8.0% | 9.8% | 6.9% | 10.2% |
| 26th-50th percentile | 7.2% | 8.3% | 7.2% | 8.0% | 6.6% | 4.5% |
| 51st-75th percentile | 6.1% | 10.1% | 6.1% | 10.9% | 6.0% | 5.5% |
| 76th-100th percentile | 4.9% | 4.7% | 4.9% | 4.7% | 5.5% | 7.3% |
| **Hospital Location by Teaching Status** | | | | | | |
| Rural | 8.9% | 14.4% | 8.9% | 16.0% | 9.8% | DS |
| Urban, non-teaching | 6.8% | 8.4% | 6.8% | 8.3% | 6.7% | 10.3% |
| Urban, Teaching | 6.2% | 8.7% | 6.2% | 9.1% | 6.2% | 5.8% |
| **Hospital Volume** | | | | | | |
| Small | 6.5% | 6.5% | 6.5% | 7.6% | 5.9% | 4.4% |
| Medium | 6.5% | 8.4% | 6.5% | 8.1% | 5.9% | 4.4% |
| Large | 6.8% | 9.5% | 6.8% | 10.0% | 7.0% | 8.0% |
| **Hospital Ownership** | | | | | | |
| Government | 9.0% | 12.6% | 9.0% | 13.3% | 8.1% | 6.1% |
| Private, non-profit | 6.0% | 8.1% | 6.0% | 8.3% | 6.0% | 6.6% |
| Private, for-profit | 8.2% | 7.8% | 8.2% | 8.2% | 7.4% | 7.5% |
| **Hospital Region** | | | | | | |
| Northeast | 5.3% | 6.9% | 5.3% | 6.8% | 4.8% | 7.4% |
| Midwest | 5.9% | 7.0% | 5.9% | 7.3% | 5.5% | 4.6% |
| South | 8.1% | 10.3% | 8.1% | 10.6% | 7.6% | 7.7% |
| West | 5.8% | 7.5% | 5.8% | 8.7% | 4.8% | 6.3% |
| **Hospital Census Division** | | | | | | |
| New England | 5.3% | 8.0% | 5.3% | 8.5% | 4.4% | 3.8% |
| Mid-Atlantic | 5.3% | 6.7% | 5.3% | 6.4% | 4.8% | 8.7% |
| East North Central | 5.9% | 7.7% | 5.9% | 7.6% | 5.6% | 4.7% |
| West North Central | 6.0% | DS | 6.0% | 5.7% | 5.0% | DS |
| South Atlantic | 7.7% | 9.6% | 7.7% | 9.5% | 7.4% | 6.0% |
| East South Central | 8.2% | 13.7% | 8.1% | 14.4% | 7.7% | DS |
| West South Central | 8.7% | 10.7% | 8.7% | 11.7% | 8.1% | 14.3% |
| Mountain | 5.8% | DS | 5.8% | 7.8% | 5.2% | 0.0% |
| Pacific | 5.8% | 7.8% | 5.8% | 8.9% | 4.6% | 8.3% |
| **Year** | | | | | | |
| 2012 | 6.9% | 11.3% | 6.9% | 10.4% | 7.2% | 4.8% |
| 2013 | 7.0% | 11.5% | 7.0% | 12.9% | 7.3% | 6.7% |
| 2014 | 6.8% | 9.2% | 6.8% | 9.5% | 7.3% | 7.8% |
| 2015 | 6.6% | 7.6% | 6.6% | 7.6% | 7.0% | 6.4% |
| 2016 | 6.6% | 8.6% | 6.6% | 9.5% | 6.3% | 9.8% |
| 2017 | 6.5% | 9.2% | 6.5% | 9.3% | 5.8% | 5.4% |
| 2018 | 6.4% | 6.7% | 6.4% | 7.2% | 5.8% | 6.4% |
| 2019 | 6.3% | 8.1% | 6.3% | 8.4% | 5.5% | 5.6% |

*** p<0.01, ** p<0.05, * p<0.1

SCD: Sickle Cell Disease

CF: Cystic Fibrosis

SMM: Severe Maternal Morbidity

DS: Data suppressed as cell counts are between 1-10

**Table S4.** Association of patient and hospital characteristics with postpartum tubal sterilization by comparison group

|  | **SCD** (N= 18,860) | | | **Non-SCD** (N= 29,803,658) | | | **Black, SCD** (N= 15,200) | | | **Black, non-SCD** (N= 4,157,700) | | | **CF** (N= 2,945) | | |
| --- | --- | --- | --- | --- | --- | --- | --- | --- | --- | --- | --- | --- | --- | --- | --- |
|  | OR | 95% CI | p-value | OR | 95% CI | p-value | OR | 95% CI | p-value | OR | 95% CI | p-value | OR | 95% CI | p-value |
| **Age, years** | 1.11 | 1.90-1.12 | <0.001 | 1.11 | 1.11 - 1.11 | <0.001 | 1.12 | 1.10 - 1.14 | <0.001 | 1.12 | 1.12 - 1.12 | <0.001 | 1.08 | 1.03 - 1.14 | 0.003 |
| **Race/Ethnicity** |  |  | 0.06 |  |  | <0.001 |  |  |  |  |  |  |  |  | – |
| White | Ref | – | – | Ref | – | – | – | – | – | – | – | – | Ref | – | – |
| Black | 1.68 | 0.88-3.21 | 0.12 | 1.10 | 1.08-1.11 | <0.001 | – | – | – | – | – | – | 2.42 | 0.53-11.09 | 0.25 |
| Hispanic | 2.19 | 1.05-4.60 | 0.04 | 1.38 | 1.35-1.41 | <0.001 | – | – | – | – | – | – |  |  | 0.99 |
| Asian or Pacific Islander | 0.53 | 0.11-2.48 | 0.42 | 0.65 | 0.63-0.66 | <0.001 | – | – | – | – | – | – | Ref |  |  |
| Native American | – | – | – | 1.11 | 1.06-1.17 | <0.001 | – | – | – | – | – | – | 1.17 | 0.34-4.06 | 0.80 |
| Other | 1.04 | 0.40-2.69 | 0.94 | 0.89 | 0.86-0.93 | <0.001 | – | – | – | – | – | – | 1.07 | 0.40-2.88 | 0.89 |
| **SMM** |  |  |  |  |  |  |  |  |  |  |  |  | – | – | – |
| No | Ref | – | – | Ref | – | – | Ref | – | – | Ref | – | – | – | – | – |
| Yes | 2.20 | 1.51-3.21 | <0.001 | 1.33 | 1.28-1.38 | <0.001 | 2.31 | 1.54-3.45 | <0.001 | 1.59 | 1.47-1.71 | <0.001 | 0.81 | 0.10-6.34 | 0.84 |
| **Delivery Mode** |  |  |  |  |  |  |  |  |  |  |  |  |  |  | – |
| Vaginal | Ref | – | – | Ref | – | – | Ref | – | – | Ref | – | – | Ref | – | – |
| Cesarean | 6.19 | 4.59-8.35 | <0.001 | 9.15 | 8.66-9.04 | <0.001 | 5.98 | 4.30-8.32 | <0.001 | 7.73 | 7.48-7.99 | <0.001 | 8.13 | 3.76-17.53 | <0.001 |
| **Median Household Income by Zip Code** |  |  | 0.01 |  |  | <0.001 |  |  | 0.02 |  |  | <0.001 |  |  | 0.27 |
| 0-25^th^ percentile | Ref | – | – | Ref | – | – | Ref | – | – | Ref | – | – | Ref | – | – |
| 26th-50^th^ percentile | 0.87 | 0.65-1.17 | 0.37 | 0.90 | 0.89-0.91 | <0.001 | 0.80 | 0.58-1.11 | 0.19 | 0.95 | 0.93- 0.97 | <0.001 | 0.41 | 0.16-1.07 | 0.07 |
| 51st-75^th^ percentile | 1.08 | 0.79-1.46 | 0.64 | 0.75 | 0.74-0.76 | <0.001 | 1.13 | 0.81-1.57 | 0.48 | 0.86 | 0.84-0.89 | <0.001 | 0.52 | 0.21-1.29 | 0.16 |
| 76^th^-100^th^ percentile | 0.48 | 0.30-0.76 | 0.002 | 0.59 | 0.58-0.60 | <0.001 | 0.46 | 0.26-0.80 | 0.007 | 0.79 | 0.75-0.83 | <0.001 | 0.70 | 0.30-1.62 | 0.40 |
| **Insurance Type** |  |  | 0.39 |  |  | <0.001 |  |  | 0.56 |  |  | <0.001 |  |  | 0.32 |
| Public | Ref | – | – | Ref | – | – | Ref | – | – | Ref | – | – | Ref | – | – |
| Private | 0.85 | 0.66-1.10 | 0.22 | 0.67 | 0.66-0.68 | <0.001 | 0.90 | 0.67-1.19 | 0.43 | 1.00 | 0.97-1.02 | 0.64 | 0.63 | 0.32-1.22 | 0.17 |
| Self-pay | 0.53 | 0.19-1.43 | 0.21 | 0.59 | 0.56- 0.61 | <0.001 | 0.50 | 0.16-1.59) | 0.24 | 0.48 | 0.44-0.52 | <0.001 | – | – | – |
| No charge, Other | 0.89 | 0.43-1.88 | 0.77 | 0.69 | 0.67-0.71 | <0.001 | 1.12 | 0.53-2.38 | 0.77 | 0.97 | 0.90-1.04 | 0.34 | 1.31 | 0.28-6.08 | 0.73 |
| **Hospital Location and Teaching Status** |  |  | 0.11 |  |  | <0.001 |  |  | 0.06 |  |  | <0.001 |  |  | 0.20 |
| Rural | Ref | – | – | Ref | – | – | Ref | – | – | Ref | – | – | Ref | – | – |
| Urban, nonteaching | 0.54 | 0.30-0.98 | 0.04 | 0.74 | 0.73-0.76 | <0.001 | 0.48 | 0.25-0.90 | 0.02 | 0.66 | 0.63-0.69 | <0.001 | 2.86 | 0.36-23.02 | 0.32 |
| Urban, teaching | 0.57 | 0.34-0.98 | 0.04 | 0.67 | 0.66-0.63 | <0.001 | 0.53 | 0.30-0.93 | 0.03 | 0.61 | 0.59-0.63 | <0.001 | 1.55 | 0.20-11.92 | 0.68 |
| **Hospital Volume** |  |  | 0.10 |  |  | <0.001 |  |  | 0.15 |  |  | <0.001 |  |  | 0.23 |
| Small | Ref | – | – | Ref | – | – | Ref | – | – | Ref | – | – | Ref | – | – |
| Medium | 1.31 | 0.84-2.04 | 0.26 | 0.99 | 0.96-1.02 | 0.37 | 1.08 | 0.68-1.73 | 0.74 | 0.99 | 0.94-1.05 | 0.83 | 0.99 | 0.25-3.96 | 0.99 |
| Large | 1.51 | 1.01-2.27 | 0.05 | 1.04 | 1.02-1.07 | 0.001 | 1.36 | 0.89-2.09 | 0.15 | 1.18 | 1.13-1.24 | <0.001 | 1.89 | 0.56-6.42 | 0.31 |
| **Hospital Ownership** |  |  | 0.001 |  |  | <0.001 |  |  | 0.002 |  |  | <0.001 |  |  | 0.94 |
| Government, nonfederal | Ref | – | – | Ref | – | – | Ref | – | – | Ref | – | – | Ref | – | – |
| Private, no-profit | 0.61 | 0.46-0.80 | <0.001 | 0.64 | 0.62-0.66 | <0.001 | 0.59 | 0.44-0.80 | 0.001 | 0.74 | 0.71-0.77 | <0.001 | 0.87 | 0.41-0.17 | 0.56 |
| Private, invest-own | 0.59 | 0.38-0.89 | 0.013 | 0.90 | 0.87-0.93 | <0.001 | 0.58 | 0.37-0.92 | 0.021 | 0.91 | 0.87-0.96 | <0.001 | 0.74 | 0.35-0.33 | 0.51 |
| **Hospital Census Region** |  |  | 0.007 |  |  | <0.001 |  |  | 0.02 |  |  | <0.001 |  |  | 0.72 |
| Northeast | Ref | – | – | Ref | – | – | Ref | – | – | Ref | – | – | Ref | – | – |
| Midwest | 1.01 | 0.67 - 1.54 | 0.95 | 1.12 | 1.09 - 1.16 | <0.001 | 1.09 | (1.64- 1.74) | 0.73 | 1.18 | (1.11 - 1.25) | <0.001 | 0.61 | 0.20-1.88 | 0.39 |
| South | 1.56 | 1.12 - 2.16 | 0.01 | 1.56 | 1.52 - 1.62 | <0.001 | 1.63 | (1.11 - 2.39) | 0.01 | 1.63 | (1.56 - 1.71) | <0.001 | 1.05 | 0.42-2.61 | 0.92 |
| West | 1.09 | 0.66 - 1.80 | 0.73 | 1.10 | 1.06 - 1.13 | <0.001 | 1.31 | (0.73 - 2.34) | 0.36 | 1.02 | (0.97 - 1.08) | 0.81 | 0.84 | 0.30-2.36 | 0.75 |
| **Hospital Census Division** |  |  | 0.02 |  |  | <0.001 |  |  | 0.04 |  |  | <0.001 |  |  | 0.31 |
| New England | Ref | – | – | Ref | – | – | Ref | – | – | Ref | – | – | Ref | – | – |
| Middle Atlantic | 0.81 | 0.39-1.72 | 0.60 | 1.01 | 0.96-1.07 | 0.73 | 0.73 | 0.31-1.73 | 0.48 | 1.08 | 0.98-1.19 | 0.12 | 2.38 | 0.27-21.16 | 0.48 |
| East North Central | 0.95 | 0.45-2.00 | 0.89 | 1.13 | 1.07-1.19 | <0.001 | 0.89 | 0.38-2.07 | 0.78 | 1.28 | 1.16-1.42 | <0.001 | 1.24 | 0.13-11.75 | 0.78 |
| West North Central | 0.51 | 0.18-1.45 | 0.20 | 1.14 | 1.07-1.21 | <0.001 | 0.64 | 0.21-1.98 | 0.44 | 1.15 | 0.99-1.34 | 0.06 | 1.16 | 0.10-13.57 | 0.44 |
| South Atlantic | 1.22 | 0.61-2.44 | 0.58 | 1.49 | 1.41-1.56 | <0.001 | 1.12 | 0.51-2.48 | 0.78 | 1.67 | 1.52-1.83 | <0.001 | 1.59 | 0.18-13.72 | 0.78 |
| East South Central | 1.82 | 0.85-3.91 | 0.12 | 1.59 | 1.49-1.70 | <0.001 | 1.80 | 0.76-4.23 | 0.18 | 1.80 | 1.62-2.00 | <0.001 | 0.63 | 0.04-9.62 | 0.18 |
| West South Central | 1.38 | 0.66-2.88 | 0.40 | 1.70 | 1.61-1.79 | <0.001 | 1.42 | 0.62-3.27 | 0.41 | 1.90 | 1.72-2.09 | <0.001 | 4.17 | 0.49-35.16 | 0.41 |
| Mountain | 0.74 | 0.23-2.37 | 0.61 | 1.10 | 1.04-1.16 | 0.001 | 0.91 | 0.27-3.07 | 0.88 | 1.20 | 1.07-1.35 | 0.002 | – | – | – |
| Pacific | 0.97 | 0.44-2.18 | 0.95 | 1.11 | 1.05-1.16 | <0.001 | 1.05 | 0.41-2.67 | 0.92 | 1.05 | 0.94-1.16 | 0.39 | 2.25 | 0.27-18.94 | 0.92 |
| **Year** |  |  | 0.15 |  |  | <0.001 |  |  | 0.26 |  |  | <0.001 |  |  | 0.72 |
| 2012 | Ref | – | – | Ref | – | – | Ref | – | – | Ref | – | – | Ref | – | – |
| 2013 | 1.02 | 0.65-1.61 | 0.93 | 1.01 | 0.97-1.06 | 0.59 | 1.28 | 0.77-2.11 | 0.34 | 1.01 | 0.94-1.08 | 0.77 | 1.43 | 0.30-6.72 | 0.34 |
| 2014 | 0.79 | 0.49-1.28 | 0.34 | 0.99 | 0.94-1.04 | 0.58 | 0.91 | 0.54-1.53 | 0.73 | 1.00 | 0.94-1.07 | 0.95 | 1.70 | 0.38-7.41 | 0.73 |
| 2015 | 0.64 | 0.39-1.05 | 0.08 | 0.96 | 0.91-1.00 | 0.06 | 0.71 | 0.41-1.23 | 0.22 | 0.98 | 0.91-1.05 | 0.52 | 1.37 | 0.32-5.95 | 0.22 |
| 2016 | 0.74 | 0.47-1.16 | 0.18 | 0.95 | 0.90-0.99 | 0.02 | 0.91 | 0.55-1.50 | 0.71 | 0.94 | 0.88-1.01 | <0.001 | 2.16 | 0.55-8.45 | 0.71 |
| 2017 | 0.79 | 0.51-1.24 | 0.31 | 0.93 | 0.88-0.98 | 0.004 | 0.89 | 0.54-1.47 | 0.65 | 0.89 | 0.83-0.96 | <0.001 | 1.15 | 0.26-5.02 | 0.65 |
| 2018 | 0.56 | 0.36-0.88 | 0.01 | 0.93 | 0.88-0.97 | 0.001 | 0.68 | 0.41-1.12 | 0.13 | 0.90 | 0.84-0.96 | <0.001 | 1.37 | 0.31-6.00 | 0.13 |
| 2019 | 0.69 | 0.43-1.10 | 0.12 | 0.90 | 0.86-0.95 | <0.001 | 0.80 | 0.47-1.35 | 0.40 | 0.87 | 0.82-0.93 | <0.001 | 1.18 | 0.25-5.51 | 0.40 |

N: Weighted count

SCD: Sickle Cell Disease

CF: Cystic Fibrosis

OR: Odds Ratio

Ref: Reference group (OR=1.0)

95% CI: 95% Confidence Interval

SMM: Severe Maternal Morbidity

**Table S5.** Sensitivity Analysis: Effect measure modification of the association of postpartum tubal sterilization and cystic fibrosis and by severe maternal morbidity

|  | **All deliveries** | |
| --- | --- | --- |
|  | aOR | 95% CI |
| **Main Effects** | | |
| Comparison Group | | |
| Non-CF | Ref | – |
| CF | 1.23 | 0.62 - 2.42 |
| SMM | | |
| No | Ref | – |
| Yes | 0.76*** | 0.72 - 0.79 |
| Delivery Mode | | |
| Vaginal | Ref | – |
| Cesarean | 8.31*** | 8.13 - 8.48 |
| Characteristics | | |
| Age, years | 1.12*** | 1.12 - 1.13 |
| Insurance Type | | |
| Public | Ref | – |
| Private | 0.50*** | 0.49 - 0.50 |
| Self-pay | 0.44*** | 0.42 - 0.46 |
| No charge, Other | 0.61*** | 0.59 - 0.63 |
| Median Household Income by Zip Code | | |
| 0-25^th^ percentile | Ref | – |
| 26th-50^th^ percentile | 0.50*** | 0.92 - 0.95 |
| 51st-75^th^ percentile | 0.55*** | 0.77 - 0.79 |
| 76^th^-100^th^ percentile | 0.61*** | 0.55 - 0.58 |
| Hospital Location and Teaching Status | | |
| Rural | Ref | – |
| Urban, nonteaching | 0.68*** | 0.66 - 0.70 |
| Urban, teaching | 0.59*** | 0.58 - 0.61 |
| Hospital Census Division | | |
| New England | Ref | – |
| Middle Atlantic | 0.96 | 0.90 - 1.03 |
| East North Central | 1.21*** | 1.14 - 1.29 |
| West North Central | 1.30*** | 1.21 - 1.39 |
| South Atlantic | 1.41*** | 1.34 - 1.50 |
| East South Central | 1.52*** | 1.41 - 1.62 |
| West South Central | 1.70*** | 1.60 - 1.80 |
| Mountain | 1.24*** | 1.17 - 1.32 |
| Pacific | 1.08** | 1.02 - 1.15 |
| Year | | |
| 2012 | Ref | – |
| 2013 | 1.00 | 0.95 - 1.05 |
| 2014 | 0.98 | 0.94 - 1.03 |
| 2015 | 0.94** | 0.90 - 0.99 |
| 2016 | 0.92*** | 0.87 - 0.96 |
| 2017 | 0.89*** | 0.85 - 0.93 |
| 2018 | 0.87*** | 0.83 - 0.91 |
| 2019 | 0.85*** | 0.81 - 0.89 |
| **Interaction** | | |
| SMM by CF | 2.71 | 0.67 - 10.98 |
| Cesarean delivery by CF | 0.87 | 0.40 - 1.91 |

Multivariate models adjusted for CF age, SMM, delivery mode, insurance type median household income by zip code, hospital location and teaching status, hospital census division, year, and the interaction of SCD and SMM, and SCD and delivery mode.

*** p<0.01, ** p<0.05, * p<0.1

aOR: Adjusted Odds Ratio

Ref: Reference group (aOR=1.0)

95% CI: 95% Confidence Interval

CF: Cystic Fibrosis

SMM: Severe Maternal Morbidity

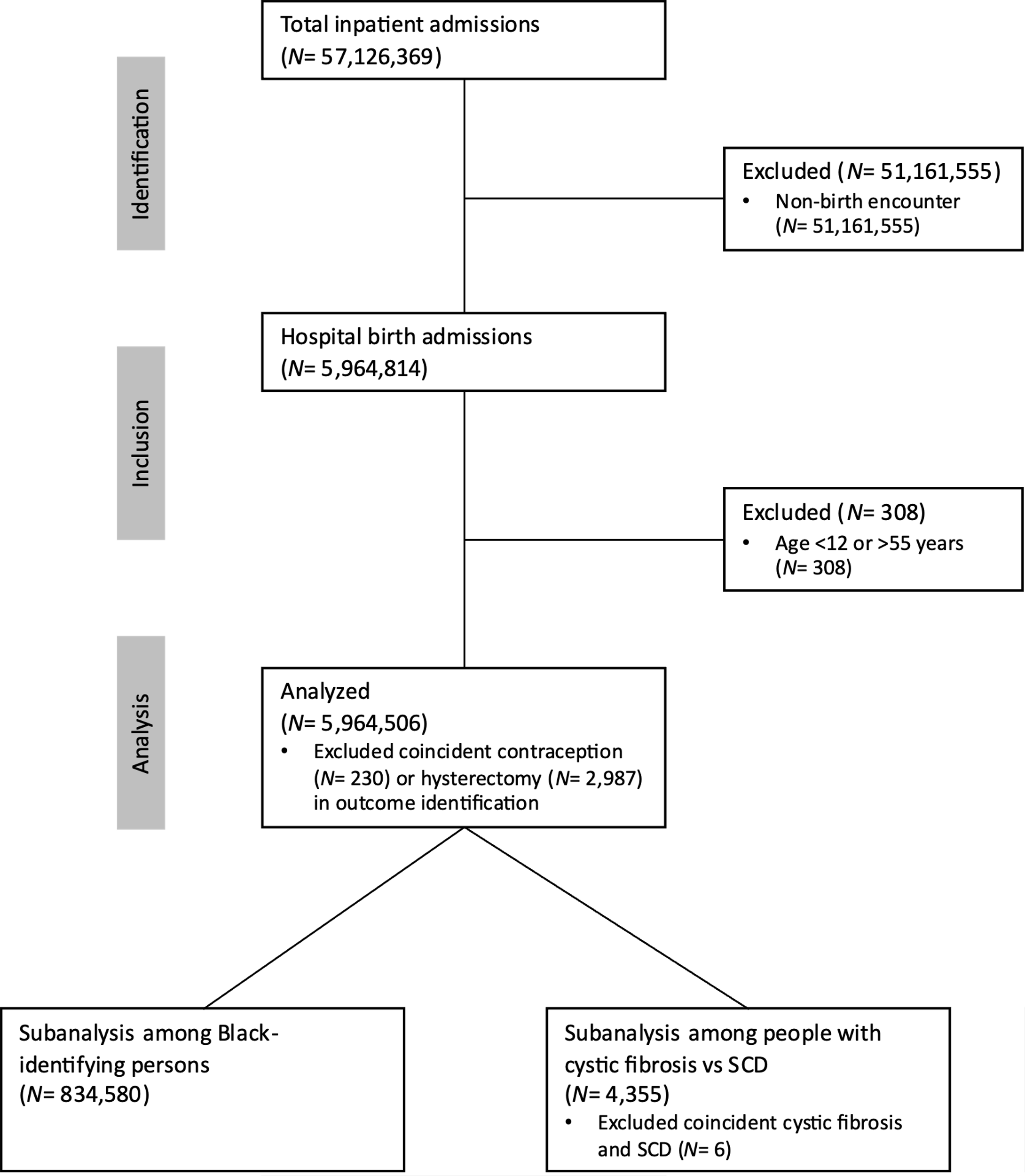

**Figure S2.** Flow diagram of study analytical sample inclusion

SCD: Sickle cell disease

**Figure S2.** Adjusted odds of postpartum tubal sterilization among sickle cell disease deliveries compared with non-sickle cell disease deliveries, stratified by hospital census division

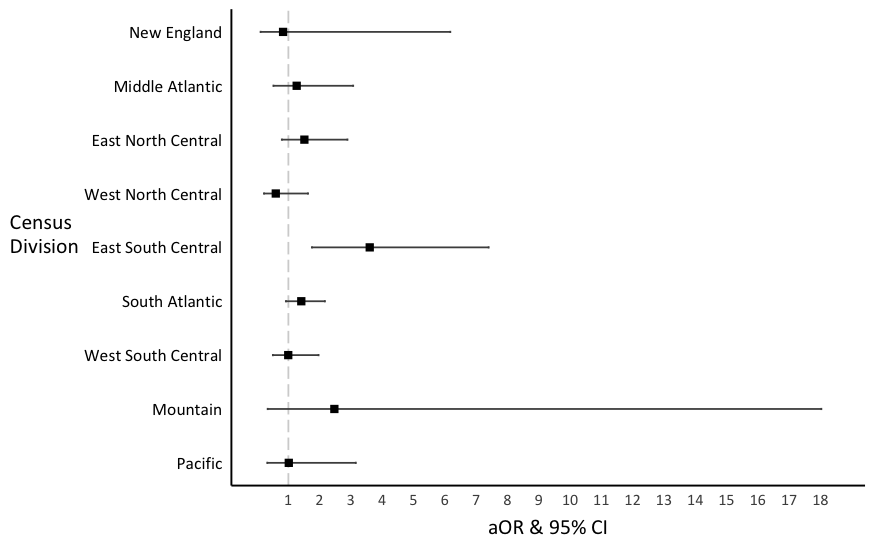

Multivariate models adjusted for SCD, age, SMM, delivery mode, insurance type median household income by zip code, hospital location and teaching status, hospital census division, year, and the interaction of SCD and SMM, and SCD and delivery mode.

aOR: Adjusted Odds Ratio

95% CI: 95% Confidence Interval
